# Heterogeneity of Diagnosis and Documentation of Post-COVID Conditions in Primary Care: A Machine Learning Analysis

**DOI:** 10.1101/2024.02.07.24302451

**Authors:** Nathaniel Hendrix, Rishi V. Parikh, Madeline Taskier, Grace Walter, Ilia Rochlin, Sharon Saydah, Emilia H. Koumans, Oscar Rincón-Guevara, David H. Rehkopf, Robert L. Phillips

## Abstract

**Background:** Post-COVID conditions (PCC) present clinicians with significant challenges due to their variable presentation.

**Objective:** To characterize patterns of PCC diagnosis in generalist primary care settings.

**Design:** Retrospective observational study

**Setting:** 519 primary care clinics around the United States who were in the American Family Cohort registry between October 1, 2021 and November 1, 2023.

**Patients:** 6,116 with diagnostic code for PCC; 5,020 with PCC and COVID-19

**Measurements:** Time between COVID-19 and PCC (U09.9) diagnostic codes; count of patients with PCC diagnostic codes per clinician; patient-specific probability of PCC diagnostic code estimated by a tree-based machine learning model trained on clinician and specific practice visited, patient demographics, and other diagnoses; performance of a natural language classifier trained on notes from 5,000 patients annotated by two physicians to indicate probable PCC.

**Results:** Of patients with diagnostic codes for PCC and COVID-19, 43.0% were diagnosed with PCC less than 4 weeks after initial recorded COVID-19 diagnostic code. Six clinicians (out of 3,845 total) made 15.4% of all PCC diagnoses. The high-performing (F1: 0.98) tree-based model showed that patient demographics, practice visited, clinician visited, and calendar date of visit were more predictive of PCC diagnostic code than any symptom. Inter-rater agreement on PCC diagnosis was moderate (Cohen’s kappa: 0.60), and performance of the natural language classifiers was poor (best F1: 0.54).

**Limitations:** Cannot validate date of COVID-19 diagnosis, as it may not reflect when disease began and could have been coded retrospectively. Few options for medically focused language models.

**Conclusion:** We identified multiple sources of heterogeneity in the documentation of PCC diagnostic codes in primary care practices after introduction of ICD-10 codes for PCC, which has created challenges for public health surveillance.

**Funding Source:** US CDC

## Introduction

Post-COVID conditions (PCC) have been challenging to study, largely due to the use of the term for a set of “potentially overlapping entities,” in the words of the US Department of Health & Human Services.(1) Understanding of PCC has evolved over time. Still, lack of detailed characterization creates difficulties for clinicians trying to treat and study the conditions.(2) The implementation of the diagnostic code for PCC (U09.9) in October 2021 gave researchers hope of a more standardized approach to diagnosing the condition.

Usage of the ICD-10 code, however, has differed substantially across clinicians, practices, and electronic health record (EHR) platforms, creating further complications for researchers. In a study from the United States Veterans Health Administration, researchers found that rates of PCC diagnosis following COVID-19 ranged from 3 to 41% across medical centers, largely due to differences in diagnostic practices.(3) Another study in the United States revealed that 35% of PCC diagnoses do not meet standards from the Centers for Disease Control and Prevention (CDC), and 60% do not meet World Health Organization (WHO) standards.(4) Meanwhile, a study from the United Kingdom found that users of different EHR platforms showed very different rates of PCC diagnosis with ICD-10 documentation.(5)

The challenge created by inconsistent usage of the PCC ICD-10 code has created demand for alternative methods of identifying patients impacted by PCC. Strategies for meeting this demand have largely been explored using machine learning methods. Zhu, et al. used a small patient cohort and symptom surveys to train a classifier on clinical notes and achieved relatively good sensitivity, albeit with a loose definition of PCC and an assumption of very high prevalence.(6) Rather than using clinical notes, other researchers have used tabular data to predict PCC. Pfaff, et al. used gradient-boosted decision trees with data contributed to the National COVID-19 Cohort Collaborative (N3C) to identify clinical and demographic features that are associated with specialty PCC clinic attendance.(7) Binka, et al. used ridge regression with administrative data from British Columbia for the same aim.(8) Among the high-prevalence test set of patients attending or diagnosed at a specialty PCC clinic, all achieved good performance, though the generalizability to patients outside that setting is unknown.

These three machine learning based studies of PCC were designed to predict whether a given patient would be diagnosed at or eligible to attend a specialty PCC clinic. Our interest, however, was in evaluating the patterns of diagnosis that exist in the generalist primary care setting. To this end, we performed a series of descriptive and machine learning-based analyses that aimed to uncover the degree and potential sources of heterogeneity in the application of the ICD-10 code for PCC among clinicians in primary care, as well as potential commonalities among patients with PCC regardless of the presence of a diagnostic code.

## Methods

### Methods overview

We combined descriptive statistical analyses with machine learning to characterize the degree of diagnostic heterogeneity of PCC within primary care and to identify potential sources thereof. We first examined the distribution of PCC diagnoses across clinicians, which allowed for characterization of clinicians’ underlying propensity to diagnose PCC. Next, we analyzed the time between the first documentation of COVID-19 and the first documentation of PCC; this is an important component of guideline-concordant diagnosis. Then, to better understand the degree to which patient and clinician factors contributed to documentation of a PCC diagnostic code, we created a gradient-boosted decision tree model trained on the patients’ other diagnoses, demographic characteristics, practice, and clinician. Finally, we trained a natural language classifier on a sample of physician-annotated clinical notes to determine whether documentation may reveal common characteristics of patients with PCC, irrespective of the presence of the PCC ICD-10 code.

### Data Source

The American Family Cohort (AFC) is a collection of EHR data derived from a registry of mostly primary care clinics across the United States.(9) The records in AFC cover the healthcare encounters between over 12,000 clinicians and approximately 8 million unique patients. Data were prospectively collected beginning in 2017 and extend in the analytic dataset to November 1, 2023 (Figure 1). The patients were of diverse ages, races, ethnicities, and geographies.

**Figure 1:**
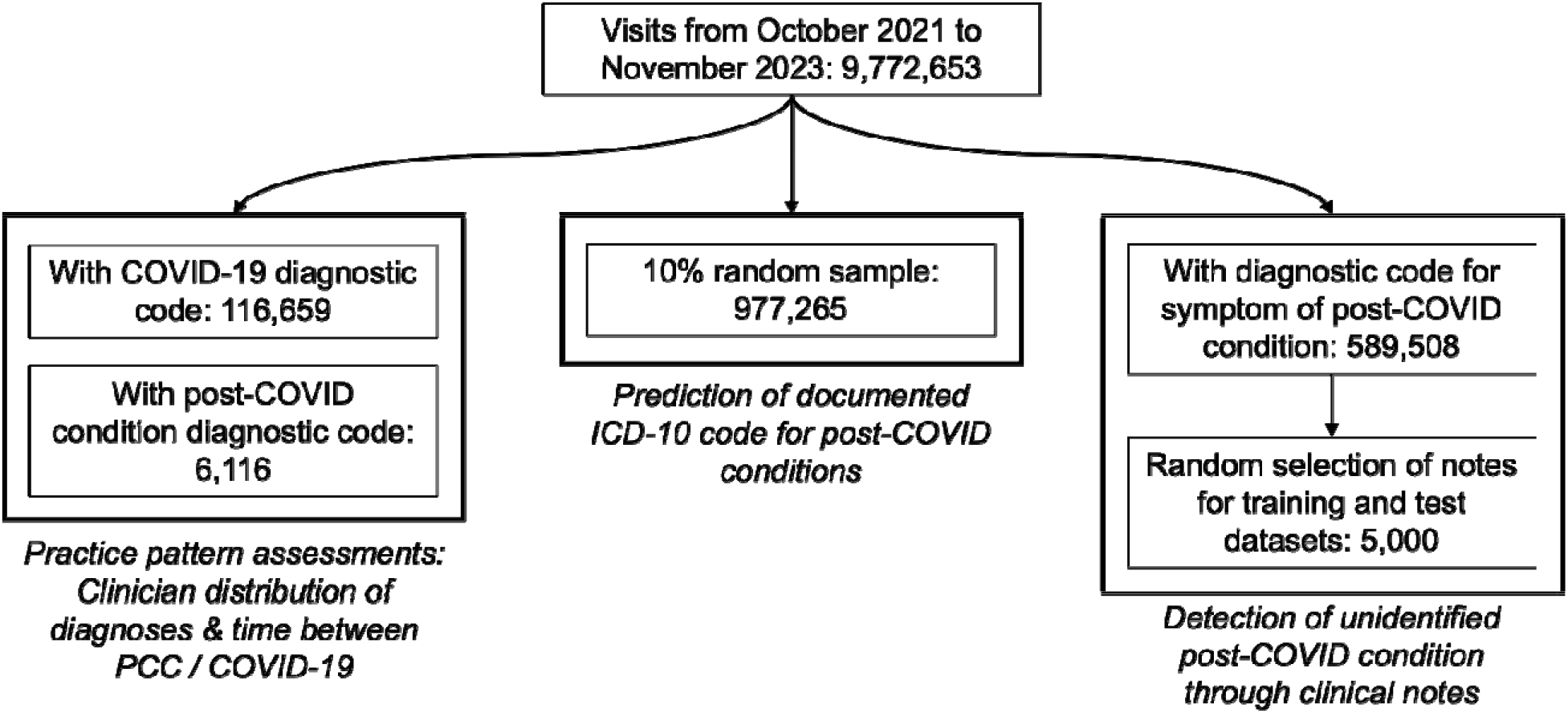
Data selection for the four included analyses. Samples were not mutually exclusive.

Approximately 20% of patients were missing data on race and ethnicity. For these patients, we used the highest probability race or ethnicity from a validated imputation based on name and census tract.(10)

### Assessment of Practice Patterns

We were interested in two proxies for understanding potential heterogeneity in application of diagnosis standards and behaviors exhibited by clinicians: first, the distribution of PCC diagnoses across clinicians; and second, the distribution of time between a patient’s first COVID-19 diagnosis and their first PCC diagnosis. Both of these analyses were conducted using the ICD-10 code U09.9 to identify patients who had been diagnosed with PCC between October 1, 2021, when the PCC ICD-10 code became available, and November 1, 2023 (Figure 1). All patients with a PCC diagnostic code were included in the descriptive analysis of how many PCC diagnoses each clinician recorded. COVID-19 was identified with the ICD-10 code U07.1 or the SNOMED code 840539006. Patients with both COVID-19 and PCC diagnostic codes were included in the analysis of time between recording the two diagnoses.

### Model of Diagnosis Code Documentation

Following the method of Pfaff, et al. (7), we developed a machine learning-based model to predict the documentation of an ICD-10 code for PCC (U09.9). As our primary interest was in examining diagnosis and documentation patterns in the context of general primary care, it was ideal that our dataset did not contain any records from specialty PCC clinics. We used as our analytic dataset a random 10% sample of visits between October 1, 2021 and November 1, 2023. This dataset included all ICD-10 codes recorded or retained within the patient’s problem list; patient demographics; date of visit; and both the practice and clinician visited. We collapsed all ICD-10 codes into the parent code: for example, the code F40.01 indicating agoraphobia would be collapsed to F40, which covers all phobic anxiety disorders. This resulted in a total of 1,596 parent codes. We used an 80% split of the 10% sample to train an extreme gradient boosted decision trees (XGBoost) model and evaluated performance on the remaining 20%.(11) Because visits labeled with a PCC diagnostic code were outnumbered by visits without, we weighted visits with a PCC diagnostic code using the inverse ratio of positive to negative labeled cases. We visualized results using Shapley values to demonstrate the influence of specific attributes on individual patients’ receipt of a PCC diagnosis code.(12)

### Sample for Natural Language Classifier

To arrive at the sample of clinical notes to train the natural language classifier we selected patient visits between October 1, 2021 and January 9, 2023 based on the recorded reason for the clinical encounter. We included only notes from visits that had a SNOMED, ICD-9, or ICD-10 diagnostic code contained in the National Library of Medicine value sets “COVID-19 Potential Signs and Symptoms” (object identifier [OID]: 2.16.840.1.113762.1.4.1223.22) or one of the ten most reported symptoms of PCC in the cross-sectional survey of PCC patients conducted by Perlis, et al.(13) (Supplemental Material I). This method was designed to capture the largest number of potential PCC patients and may have missed patients who presented with less common symptoms.

Next, we included only patient notes having a length of at least 100 characters to eliminate most of the uninformative notes and extraneous data that could be mistakenly included in the clinical notes section. We concatenated all notes for each unique combination of patient ID and visit date for the visits identified in the first step of the sampling process.

Finally, we randomly selected 5,000 clinical notes from unique patients among the subset of all notes meeting our criteria. Each note was from a single visit. This sample size is based on the sample size of a similar study that achieved good model performance.(14) Many notes contained formatting marks, which we cleaned using regular expression-based functions. This was important not only for readability by the physicians who later tagged the documents, but also for avoiding training the NLP model on formatting marks – for example, identifying metadata or fonts from particular facilities.

### Natural Language Classifier Development

Two family physicians (MT and GW) each reviewed 2,750 different notes. Both reviewed an overlapping set of 500 notes, which we used for calculating Cohen’s kappa for inter-rater reliability. The physicians used the criteria defined by the CDC to identify patients who were eligible for a PCC diagnosis.(15) Because they were using clinical notes from single visits and no structured data from patients’ charts, they also required attribution of the symptoms to a prior COVID-19 illness within the note. Within the overlapping subset, disagreements in classification were resolved by a third reviewer (NH).

We used three different NLP-based classifiers of increasing complexity: a tree ensemble model, a recurrent neural network (RNN), and a Transformer-based model. We trained each model on a 70% sample (i.e., 3,500 of 5,000 notes) and assigned the remainder to the test dataset.

For the first model, we trained an XGBoost model on term frequency-inverse document frequency (TF-IDF) data – a form of regularized bag-of-words model.(11,16) XGBoost is a tree ensemble method that has proven competitive with deep learning methods on many types of sparse tabular data, including text.(17)

For the second model, we employed a long short-term memory (LSTM) model.(18,19) Unlike bag-of-words approaches, which treat text as a static input with no consideration of word order or context, a LSTM RNN model can integrate the sequence of words and their dependencies into its predictions. We composed our model with an Embedding layer, LSTM units, and a final Dense layer, which we trained using binary cross-entropy loss and the Adam optimizer.

The third model was a transformer pre-trained on deidentified clinical notes. Because it had shown superior performance on clinical classification tasks over general transformer models, we used BioClinicalBERT as the basis for our transformer-based classifier.(20) A caveat of the transformer architecture is that the model length input is limited. In the case of BioClinicalBERT, it is limited to 128 tokens (roughly equivalent to 100 words). Therefore, we separated notes into texts of suitable length prior to processing in the transformer model. This meant, however, that the model produced multiple predictions for each note, and we had to aggregate them. We tested both the mean and maximum of predictions for each note and used the best performing aggregation method.

Because only 1% of the notes related to PCC, we augmented presumptive PCC notes and under-sampled other notes. We first applied synthetic text augmentation to the positive cases using a WordNet-based synonym augmenter, which duplicates notes and randomly replaces words with synonyms, effectively increasing the variety of positive case samples.(21) We then employed random undersampling, in which a significant portion of the negative cases was randomly dropped, thereby reducing the disparity between positive and negative instances in the dataset. With random sampling, we ensured that the ratio of negative to positive cases did not exceed 5:1.

We used the Optuna package in Python to optimize hyperparameters in the RNN and transformer models.(22) In each case, we used evaluation loss on a 30% test dataset as the cost function over 50 trials. Our primary outcome of interest was area under the receiver operating characteristic curve (AUC), and positive / negative predictive value at the optimal threshold, as determined by maximization of the F1 score. We calculated confidence intervals for each model’s performance metrics by using a bootstrap with 1,000 repetitions.

### Software

Descriptive analyses were conducted in R, version 4.2, while modeling was conducted in Python 3.7. We used the xgboost package for prediction of PCC diagnostic code documentation.(11) We used the scikit-learn(23) and xgboost packages for the TF-IDF analyses; TensorFlow for the RNN(24); and HuggingFace Transformers for the transformer.(25)

## Results

### Dataset Characteristics

The AFC contained 9,722,653 visits conducted by 3,845 clinicians at 519 practices with 4,724,507 unique patients from October 1, 2021, to November 1, 2023. Among these, 116,659 patients had a diagnostic code for COVID-19 and 6,116 had a diagnostic code for PCC. A total of 5,020 individuals had diagnostic codes for both COVID-19 and PCC: 1,096 (18%) patients with a PCC diagnostic code did not have a diagnostic code for COVID-19.

### Patterns of Documentation for the PCC Diagnostic Code

Of the 3,845 clinicians, 973 (25.3%) documented a PCC diagnostic code for at least one patient. The greatest number of PCC diagnoses took place in January and February of 2022, following the 2021 surge of infections driven by the emergence of the Omicron variant of SARS-CoV-2 (Supplemental Material II). The distribution of PCC diagnoses was highly right-skewed (Figure 2). A substantial share of the clinicians who diagnosed any PCC, 331 (34.0%), diagnosed only one patient with PCC. Six clinicians had over 100 patients with PCC, and the maximum number of PCC diagnoses for a single provider was 224. These six clinicians, all of whom practice in different states without any public indication of working at a specialty PCC clinic, accounted for 15.6% (957 out of 6,116) of all PCC diagnoses documented with diagnostic codes. Seven clinicians diagnosed more than 10% of their patients with PCC, and 35 diagnosed more than 5%.

**Figure 2:**
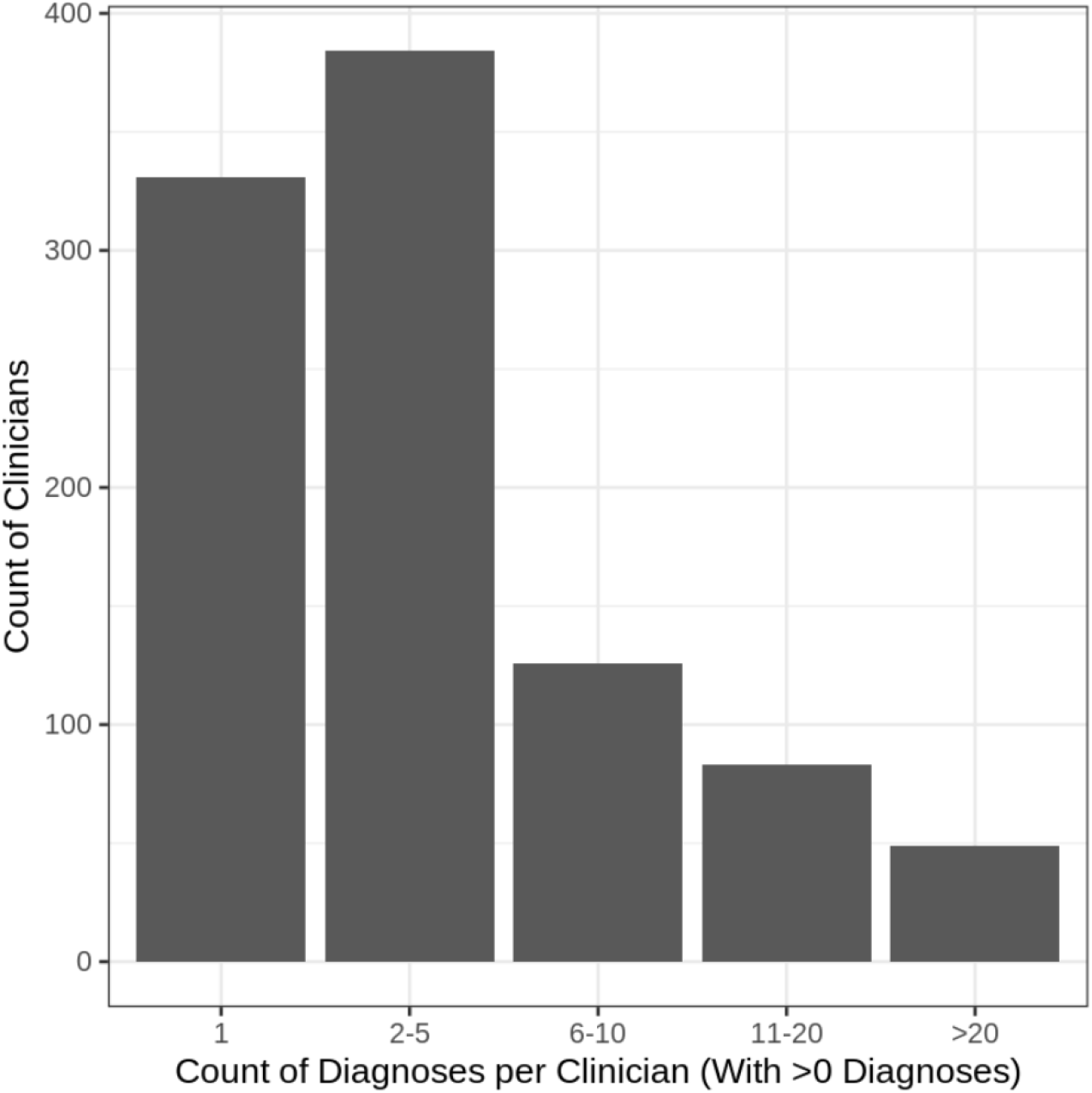
Diagnoses made for each of the 973 (out of 3,845 total) clinicians with at least one PCC diagnosis made from October 1, 2021 – November 1, 2023.

### Time between COVID-19 and PCC diagnoses

Among patients with diagnostic codes for both COVID-19 and PCC, 295 (5.8%) had a PCC diagnosis recorded before their first documented diagnosis of COVID-19. An additional 3,078 (61.3%) individuals received their first PCC diagnosis less than 12 weeks after their first COVID-19 diagnosis, and 2,158 (43.0%) were diagnosed with COVID-19 and PCC less than 4 weeks apart. These thresholds represent the timing of diagnosis specified by the WHO and CDC, respectively. The mean time between first COVID-19 and first PCC diagnosis among those with both was 127 days, and the median time was 30 days.

### Predictive Model of Diagnostic Code Documentation

The XGBoost model trained on documentation of a diagnostic code for PCC achieved excellent performance. Its overall accuracy was 99.7% (95% confidence interval [CI]: 99.7 to 99.8%), its weighted F1 score was 0.982 (95% CI: 0.982 to 0.983), and its AUC was 0.711 (95% CI: 0.679 to 0.741). Overall, patient demographics were more predictive of the presence of a PCC diagnostic code than any recorded diagnosis and clinical features (Figure 3). Older age, female gender, and non-Hispanic white race/ethnicity were all associated with higher rates of PCC documentation. Similarly, calendar date and the practice and clinician visited were more important than any recorded diagnoses. Force plots for four example patients (Supplemental Material III) show similar patterns.

**Figure 3:**
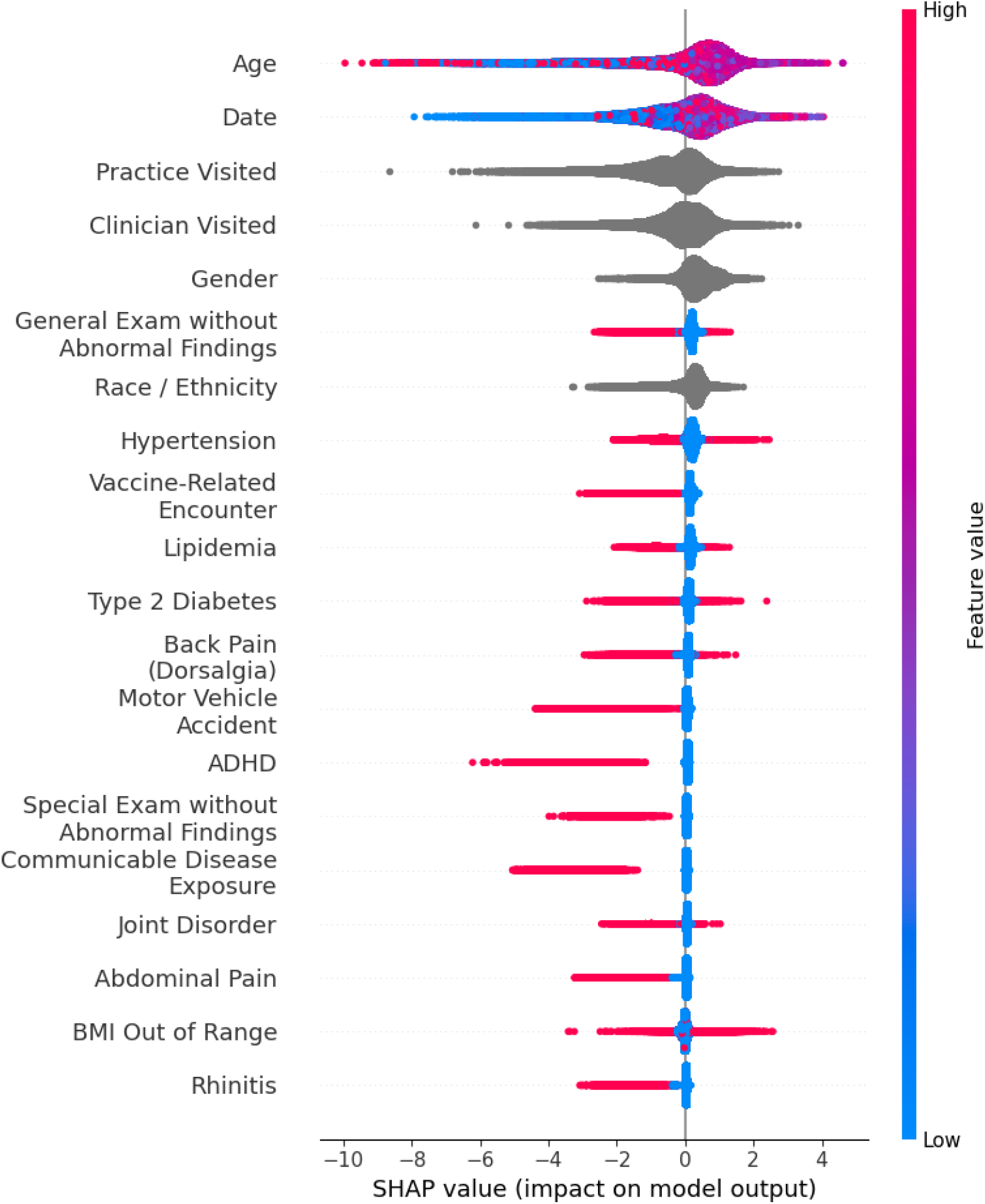
Shapley values for the model predicting documentation of an ICD-10 code (U09.9) for post-COVID conditions. Feature importance across patients is represented on the vertical axis (higher values on top), while influence on individual patients is represented by the jittered points along the horizontal axes. Feature value is indicated by color; for instance, low age is blue while higher age is red, and the presence of a given clinical condition is indicated by red. Gray indicates nominal categorical variables with more than two potential values.

### Natural Language Classifier Performance

In the annotated sample, 50 of the 5,000 notes were related to PCC. Interrater reliability calculations between the two physician reviewers showed a Cohen’s Kappa of 0.6 for PCC, which indicates moderate agreement beyond chance.

The models had low accuracy at identifying patients with PCC. The model AUCs were 0.76, 0.58, and 0.45 for the TF-IDF, RNN, and transformer models, respectively (Table 1, Supplemental Material IV). Only the TF-IDF model was better than chance. Confusion matrices (Supplemental Material V) showed that the RNN and transformer models achieved their best performance by classifying all individuals as not having PCC.

**Table 1:** Performance characteristics of the three natural language classifiers (with 95% confidence intervals). Performance is based on physician review of notes within a 30% set-aside test set. AUC = area under the curve; RNN = recurrent neural network; TF-IDF = term frequency-inverse document frequency (subsequently used in a tree-based classifier).

|  | AUC | F1 Score | Positive predictive value | Negative predictive value | Sensitivity | Specificity |
| --- | --- | --- | --- | --- | --- | --- |
| TF-IDF | 0.76 (0.54 – 0.93) | 0.54 (0.32 – 0.65) | 0.41 (0.23 – 0.56) | 0.94 (0.87 – 0.99) | 78% (50 – 92%) | 77% (68 – 85%) |
| RNN | 0.58 (0.36 – 0.85) | 0.37 (0.06 – 0.48) | 0.23 (0.08 – 0.67) | 0.94 (0.77 – 0.99) | 39% (5 – 43%) | 89% (65 – 97%) |
| Transformer | 0.45 (0.22 – 0.67) | 0.32 (0.08 – 0.47) | 0.32 (0.09 – 0.50) | 0.86 (0.77 – 0.93) | 33% (8 – 50%) | 85% (77 – 92%) |

## Discussion

Our results revealed substantial heterogeneity in the diagnostic behavior of clinicians in primary care in the 15 months following the introduction of the ICD-10 code for PCC. This suggests – but does not prove – that interrater reliability of PCC diagnosis among clinicians in primary care is low. While a majority of clinicians in our dataset (75.7%) did not record a single diagnostic code for PCC, others applied the diagnosis very widely. Our data also showed that a majority (61.3%) of PCC diagnoses did not seem to meet the WHO criteria of at least twelve weeks between COVID-19 and PCC; nearly half (43.0%) did not meet the CDC definition of a minimum four weeks. Furthermore, our machine learning analyses revealed that patient demographics, practice visited, and clinician visited were more predictive of a PCC diagnostic code than most recorded clinical factors. Three NLP models could not meaningfully identify PCC from the text of clinical notes, suggesting that patient heterogeneity may also be high, in addition to clinician heterogeneity.

PCC is a collection of symptoms that may commonly occur together but with wide variation. The CDC’s PCC symptom collection originally contained more than 1500 symptoms. which may mean that we have not sufficiently winnowed the most precise collection of symptoms to define PCC. Clusters of patients may have different PCC clusters with little overlap between them, thereby producing variation in their presentation at primary care. In practice, primary care providers such as those represented in AFC have reported difficulties diagnosing and treating PCC patients in the face of often ambiguous definitions and standards.(26) The annotations of clinical notes from the two physicians demonstrated this issue: the level of agreement was only moderately above what would be expected by chance alone.

Our study revealed results that broadly accord with the findings of other studies. Prior research found low concordance between the ICD-10 code for PCC and the clinical criteria for the disease, (4) and a right-skewed distribution of PCC diagnoses across clinicians.(5) Prior machine learning-based studies that focused on counterfactual probability of diagnosis had patients visited specialty PCC clinics all found different estimates of the mean risk of PCC following COVID-19, ranging from approximately 20% in Binka, et al. to over 40% in Pfaff, et al.(7,8) This mirrors an even wider difference in estimates derived from prospective patient surveys, which range from 4.5% to 89% --a difference that has been attributed to heterogeneous case definitions.(27) Our study’s results also agreed with prior research that found PCC diagnoses demographically cluster among non-Hispanic white female patients.(28)

Our results point to unmet needs for clinicians, patients, and researchers. Clinicians in general primary care settings have played an important role in identifying and managing PCC, yet report feeling poorly equipped to treat it holistically.(26) In practice, clinicians may choose to focus on individual symptoms rather than the collective condition and to treat PCC as a diagnosis of exclusion.(29,30) Cau, et al. suggested that artificial intelligence could have a role in supporting PCC treatment across practices.(31) Meanwhile, patients face many unmet needs as their symptoms persist.(32) Researchers, too, face challenges in identifying patients with PCC in observational databases, making it difficult to characterize these conditions’ prevalence, trajectory, and treatment.(33)

Our study had a number of limitations. First, the physician annotators only had access to notes from a single visit, nor did they have access to any diagnostic codes, vital signs, or labs. They were therefore limited in the amount of context they could use in their ascertainments. A second limitation is that the Wordnet model we used was not specialized for medical text. We are not aware of any clinically focused Wordnet dictionaries that we could have used in its place and did not find any gross errors on inspection. Third, our choice of BioClinicalBERT meant that we used a transformer model capable of accepting only a relatively small amount of text at a time; another transformer model may have performed better. Fifth, the dataset was highly imbalanced, with PCC diagnoses included in a relatively small portion of visits. Finally, we cannot validate the dates documented for any diagnosis entered into an EHR. Thus, if a patient were diagnosed with COVID-19 in a setting not captured within our data (e.g., inpatient setting, emergency department), the accuracy of the date of first documented diagnosis recorded in the EHR depended upon the clinician who documented it. Similarly, the documented date of diagnosis may be delayed if the patient did not seek care upon initially testing positive outside of a clinical encounter. Thus, date associated with diagnosis code may not reflect when disease began and could be a “retrospective” code reflecting prior illness not captured previously in the EMR.

This study was strengthened by the use of the real-world data that captured the experience of diagnosing PCC in a diverse set of primary care settings. Our use of architectures validated on classifying COVID-19 diagnoses gave credence to our findings that even highly sophisticated NLP models struggle to accurately identify PCC patients from notes alone. Moderate inter-rater reliability metrics between the two physician annotators also highlighted the challenges of identifying PCC, even for expert reviewers.

Given the heterogeneity around diagnosis of PCC among the included providers, one area for future research may be the development of explicitly counterfactual approaches to identification of PCC cohorts. This may involve the development of provider-specific models of diagnosis that indicate the likelihood that they would diagnose a patient with a given presentation.

Researchers could then use a single diagnostic model across an entire population to standardize diagnosis across providers.

## Conclusion

This research points to multiple sources of heterogeneity affecting the documentation of PCC. Wide variation in diagnostic and documentation practices are likely due to lack of definitive diagnostic criteria for this syndrome in the period of observation, making it difficult for sophisticated natural language classifiers to reliably detect. As guidance for PCC diagnosis stabilizes and frontline clinicians are more aware of the ICD-10 code diagnostic code, its use for public health surveillance may grow. It will be useful to continue to monitor trends in use of the PCC ICD-10 diagnostic code and to simultaneously use NLP or other methods to understand the symptoms most often associated in primary care, where most people present for undifferentiated symptoms. Lack of clear diagnostic criteria with face validity in primary care may continue to contribute to inconsistent documentation practices and barriers to effective care for patients with PCC.

## Data Availability

Data may be available for IRB-approved research subject to approval of the American Board of Family Medicine Research Governance Board.

