## Supplemental for "Heterogeneity of Diagnosis and Documentation of Post-COVID Conditions in Primary Care: A Machine Learning Analysis"

I. *Symptoms included in identification of candidate cohort. Object identifiers are from the National Library of Medicine Value Set Authority Center.*

| **Symptom** | **Object Identifier(s)** |
| --- | --- |
| Shortness of breath | 2.16.840.1.113762.1.4.1182.47 |
| Fatigue | 2.16.840.1.113762.1.4.1146.862 and 2.16.840.1.113762.1.4.1146.860 |
| Headache | 2.16.840.1.113883.17.4077.3.1027 |
| Loss of smell | 2.16.840.1.113762.1.4.1146.1202 and 2.16.840.1.113762.1.4.1146.1201 |
| Brain fog | 2.16.840.1.113762.1.4.1222.1377 |
| Poor memory | 2.16.840.1.113883.3.3616.200.110.102.3221 and 2.16.840.1.113883.3.3616.200.110.102.6310 |
| Dizziness | 2.16.840.1.113883.3.3616.200.110.102.3223 and 2.16.840.1.113883.3.3616.200.110.102.6315 |
| Depressed mood | 2.16.840.1.113883.3.600.145 |
| Anxious mood | 2.16.840.1.113762.1.4.1021.94 |
| Sleep disruption | 2.16.840.1.113762.1.4.1222.1318 |

II. *Monthly diagnoses of PCC in the time since the PCC ICD-10 code became available October 2021 – October 2023.*

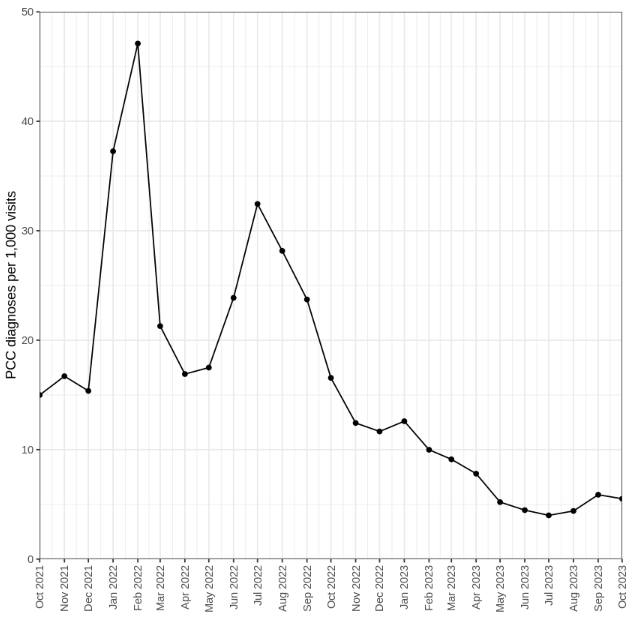

III. *Force plots for randomly selected negative- (no PCC diagnostic code) and positive- (PCC diagnostic code) labeled visits.*

Negative examples

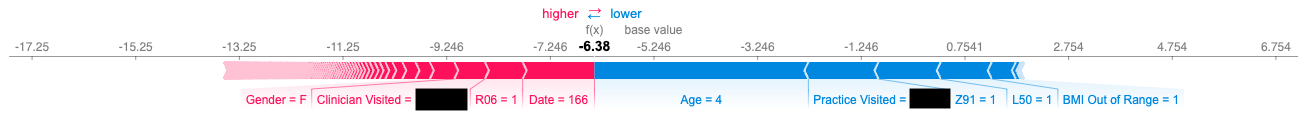

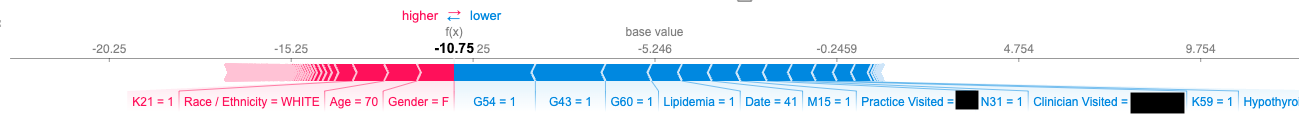

Positive examples

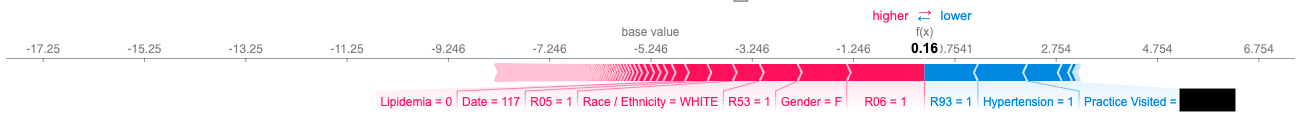

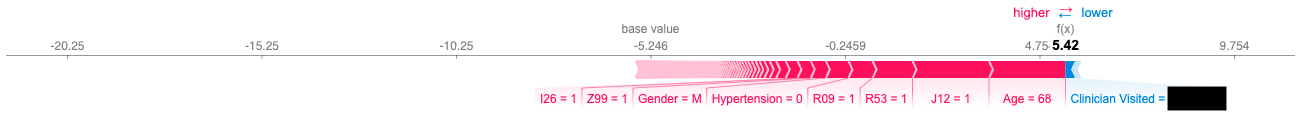

Notes: XGBoost captures many non-linearities and interactions in data. As such, the impact of a given parameter differs in amount and sometimes even sign across patients. Some data were redacted to ensure patient privacy. The date variable is the number of days since October 1, 2021.

V. *Receiver operating characteristic curve for the three natural language classifiers trained to identify PCC. AUC = area under the curve; RNN = recurrent neural network; TF-IDF = term frequency-inverse document frequency (subsequently used in a tree-based classifier)*

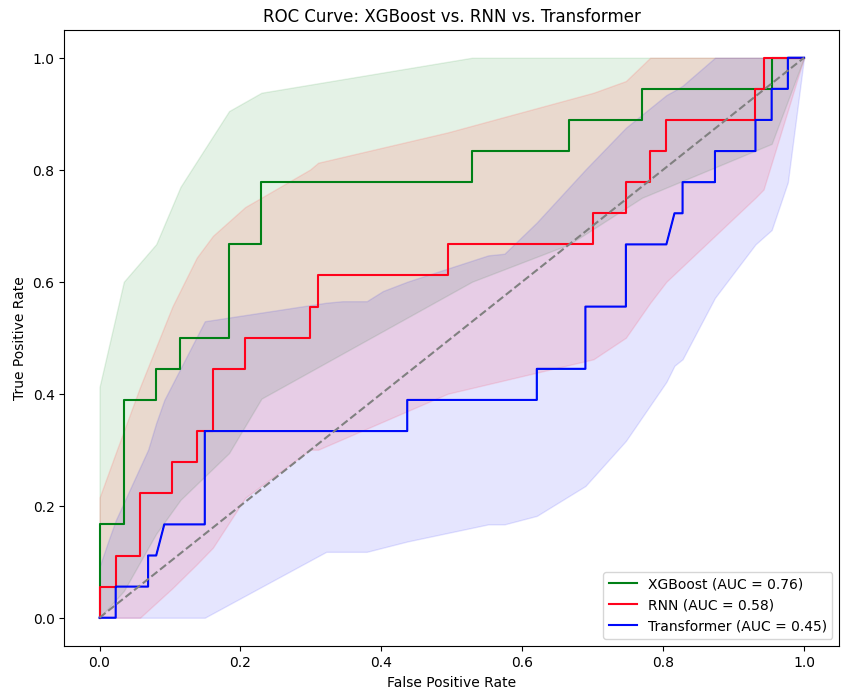

VI. *Confusion matrices for the three NLP models*

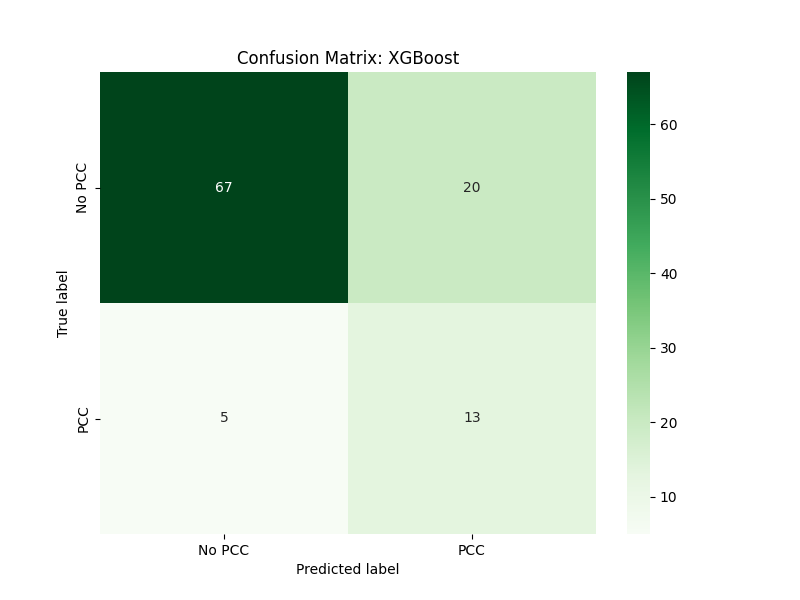

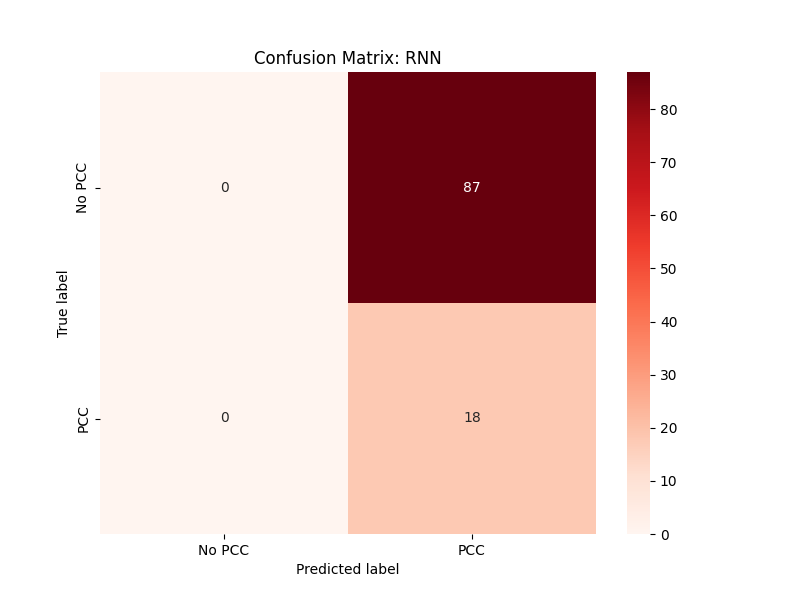

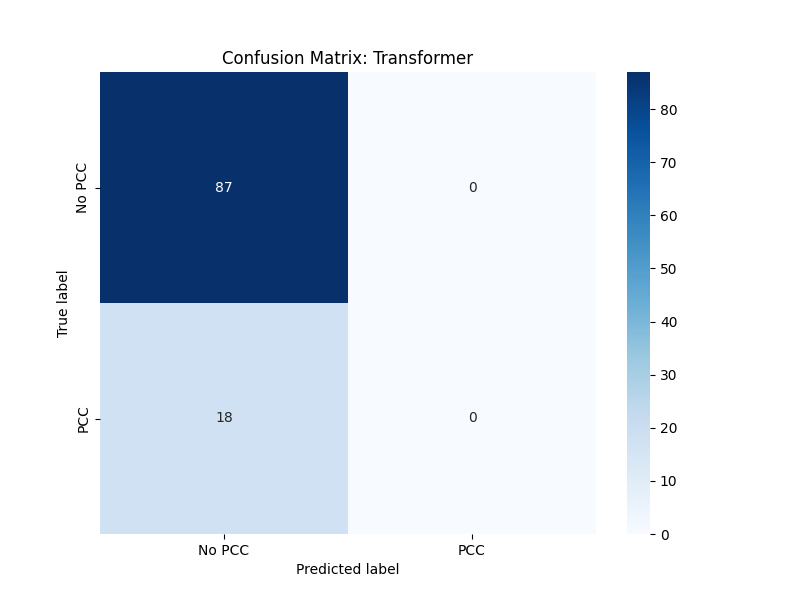
